# Toward Digital Phenotypes of Early Childhood Mental Health via Unsupervised and Supervised Machine Learning

**DOI:** 10.1101/2023.02.24.23286417

**Authors:** Bryn C. Loftness, Donna M. Rizzo, Julia Halvorson-Phelan, Aisling O’Leary, Shania Prytherch, Carter Bradshaw, Anna Jane Brown, Nick Cheney, Ellen W. McGinnis, Ryan S. McGinnis

## Abstract

Childhood mental health disorders such as anxiety, depression, and ADHD are commonly-occurring and often go undetected into adolescence or adulthood. This can lead to detrimental impacts on long-term wellbeing and quality of life. Current parent-report assessments for pre-school aged children are often biased, and thus increase the need for objective mental health screening tools. Leveraging digital tools to identify the behavioral signature of childhood mental disorders may enable increased intervention at the time with the highest chance of long-term impact. We present data from 84 participants (4-8 years old, 50% diagnosed with anxiety, depression, and/or ADHD) collected during a battery of mood induction tasks using the ChAMP System. Unsupervised Kohonen Self-Organizing Maps (SOM) constructed from movement and audio features indicate that age did not tend to explain clusters as consistently as gender within task-specific and cross-task SOMs. Symptom prevalence and diagnostic status also showed some evidence of clustering. Case studies suggest that high impairment (>80th percentile symptom counts) and diagnostic subtypes (ADHD-Combined) may account for most behaviorally distinct children. Based on this same dataset, we also present results from supervised modeling for the binary classification of diagnoses. Our top performing models yield moderate but promising results (ROC AUC .6-.82, TPR .36-.71, Accuracy .62-.86) on par with our previous efforts for isolated behavioral tasks. Enhancing features, tuning model parameters, and incorporating additional wearable sensor data will continue to enable the rapid progression towards the discovery of digital phenotypes of childhood mental health.

**Clinical Relevance:** This work advances the use of wearables for detecting childhood mental health disorders.

## I. INTRODUCTION

Mental health disorders in early childhood are prevalent, disruptive, and can negatively impact development if left unaddressed [1]-[3]. Children cannot reliably report their abstract emotions [4], and guardian reports are often biased, leading to challenges in detection of childhood mental health conditions [5], [6]. There is a persisting need for objective measures to supplement current childhood mental health assessments, increasing reliability and sensitivity. Our prior work lays the groundwork for these efforts by introducing a free open-access researcher-accessible digital phenotyping application, ChAMP, and a corresponding battery of behavioral mood induction tasks for assessing childhood mental health [7]-[15] (see Fig. 1). While our preliminary work provides compelling evidence that data from individual mood induction tasks can be used to detect children with internalizing disorders (e.g., anxiety), these results need to be replicated in a larger cohort and extended to consider data from the entire task battery. Moreover, in line with the Research Domain Criteria (RDoC), there is also a need to explore data from the task battery for latent groupings that may be indicative of internalizing or externalizing impairment, rather than specific diagnoses.

**Figure 1.**
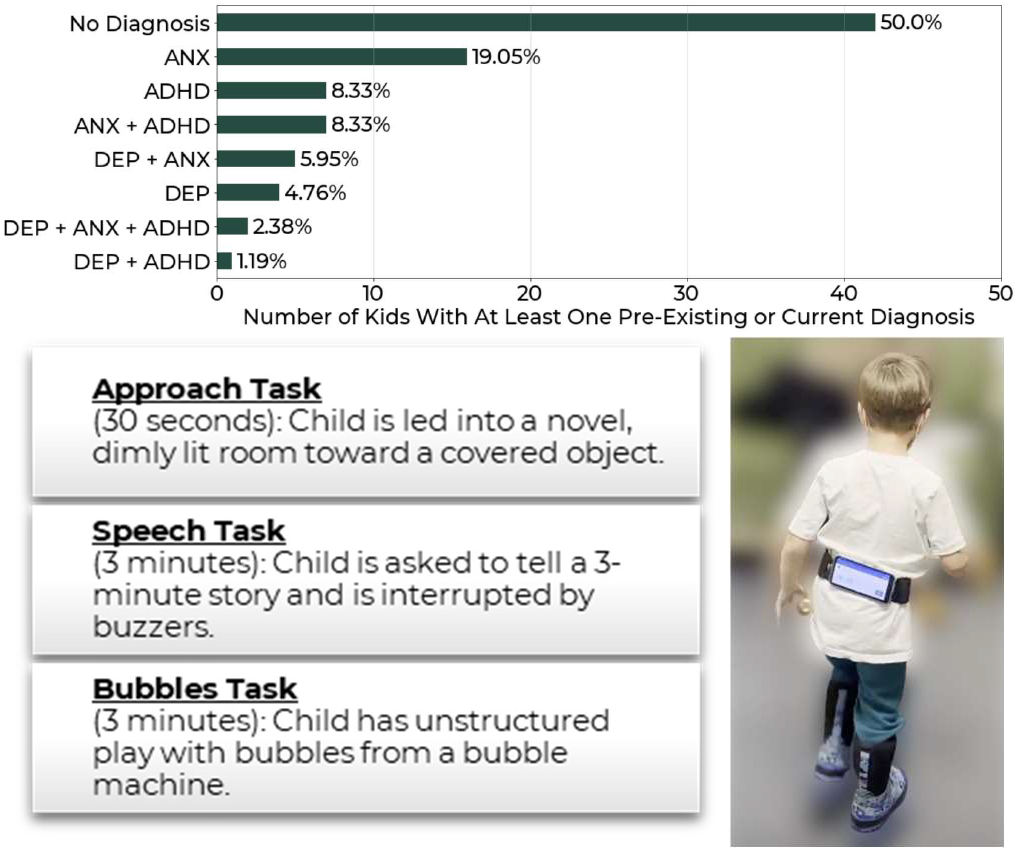
Cohort by diagnostic status. DEP = Depression, ANX = Anxiety, ADHD = ADHD (**top**). Descriptions of ChAMP app tasks included in the assessment battery (**bottom left**). Photo of a child wearing the ChAMP app waistband (**bottom right**).

Herein, we extend our prior work by exploring the use of unsupervised clustering for identifying latent groupings within ChAMP behavioral assessment battery data and supervised learning to determine if we can build classifiers for detecting children with ADHD, anxiety, or depression. These efforts inform the potential for future clustering and predictive modeling in child mental health using robust feature sets of wearable sensor-derived data.

## II. Methods

For this analysis, we consider data from the Kiddie Internalizing Disorder (KID) Study [14], [15]. Children in the KID Study participate in an administrator-led, RDoC-motivated battery of behavioral mood induction tasks that relate to a variety of internalizing and externalizing disorders [16], [17]. During this assessment, the child wears an array of wearable sensors and a smartphone, equipped with the ChAMP app [15] which record physiological, behavioral, and vocal data throughout the tasks. Simultaneously and separately, the guardian is guided through a semi-structured diagnostic interview and completes mental health surveys for themselves and the child. Child diagnosis is determined through a gold-standard consensus coding technique (summary in Fig. 1). Of the 95 participants enrolled in this on-going study, we consider data from 84 who have complete behavioral task and diagnostic data at the time of analysis (age in months: 81.78 +/-14.64, gender: 51 M, 33 F).

We focus on data collected via the ChAMP app (Fig. 1, right) during the behavioral task battery (Fig. 1, left). The app collects three-axis gyroscope and accelerometer data during the Approach [8] and Bubbles [7] tasks, and audio data during the Speech task [9]. Signal processing and feature extraction are described in detail in [15]. Briefly, accelerometer and gyroscope signal vector magnitudes are computed and used to inform calculation of signal features which are extracted from theory-driven temporal phases within the Approach and Bubbles tasks. Similarly, a voice activity detector is applied to the audio signal before extracting vocal features from theory-driven temporal phases within the Speech task. This process yields 617 features from the three behavioral tasks for analysis.

Univariate difference testing is used to reduce the feature set to just those features which show at least one significant difference between diagnostic groups (anxiety, depression, or ADHD vs. not anxiety, depression, or ADHD). This process yielded 24, 51, and 25 features from the Approach, Speech, and Bubbles tasks, respectively (100 total).

### A. Unsupervised Clustering

Kohonen Self-Organizing Maps (SOMs) [18] were used to discover latent clusters within features derived from the ChAMP app data for each behavioral task individually and all tasks together. Trends in vectors of individual behavioral signatures were detected using MiniSOM [19]. The SOMs underwent hyperparameter tuning to identify representations that showed cluster differentiation, and then characteristics (e.g., diagnosis) were overlayed on the SOMs after the random weight seeds were selected. All SOMs are 500×500 neurons with a gaussian neighborhood function. Each of the SOM weights are consistent across the trained SOMs visualized in this work. The cross-task, Approach, and Bubbles SOMs all have learning rates of .9, while the Speech task SOM has a learning rate of .85. The cross-task, Approach, Speech, and Bubbles SOMs use neighborhood sizes of 60, 60, 50, and 50, and unified distance matrix (U-matrix) [20] minimum grid gradients of .35, .16, .34, and .2, respectively.

### B. Supervised Learning for Mental Health Diagnosis

To assess the potential for detecting children with ADHD, anxiety, or depression, we train binary classification models. Models were trained to classify children with depression, anxiety, or ADHD *vs* children who were not diagnosed with each respective disorder based on age, gender, and the ChAMP system’s reduced feature set. Performance of Gaussian Naïve Bayes (NB), Random Forest (RF), Support Vector Machine with Stochastic Gradient Descent Booster (SVM), Decision Tree (DT), and Logistic Regression (LR) models with default hyperparameters were evaluated using 2-fold shuffled cross-validation. Performance of each classifier was characterized by area under the receiver operating characteristic curve (ROC AUC), false positive rate (FPR), true positive rate (TPR), and Mean Accuracy across the two folds’ test cases.

## III. Results

### A. Unsupervised Clustering: Behavioral Signatures

Results of the unsupervised clustering highlight each child’s unique movement and vocal signatures during the mood induction tasks, with some exhibiting more unique behaviors than others (e.g., bottom left of Fig. 2). Differences in behavioral signatures can be identified in the visualized U-matrices, which represent the Euclidean distance between neuron feature vectors within the SOM and further by proximity within the boundaries of their shaded regions (*i.e*., proximity within a boundary represents individuals with similar feature vectors).

**Figure 2.**
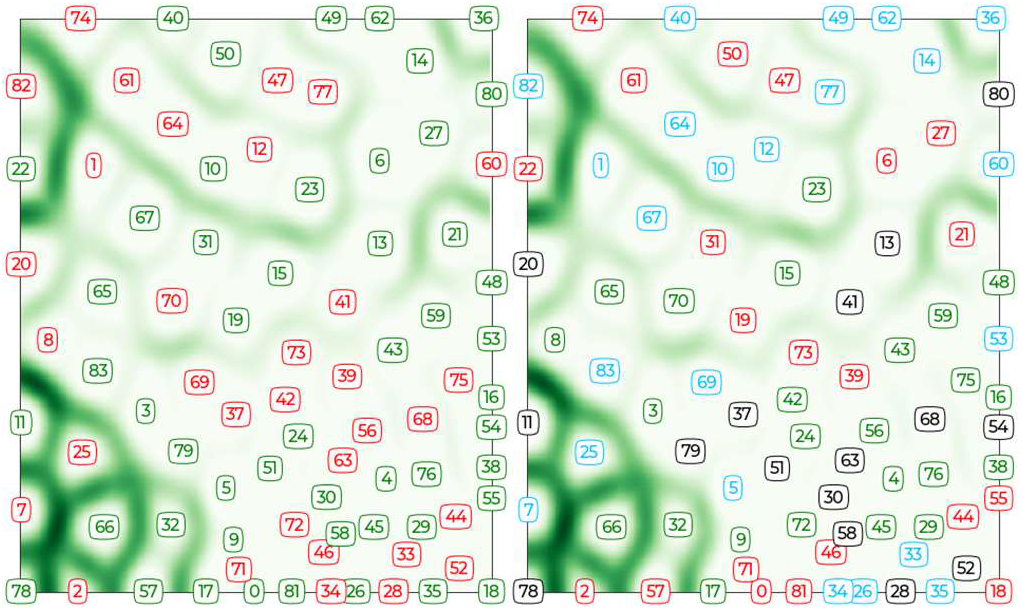
SOMs of the Approach task encoded by gender (**left**, red-female, green-male) and by age in years (**right**, 4 and 5-red, 6-green, 7-blue, 8-black).

Despite the overall evidence of individuality, the Approach task SOM representations of Fig. 2 indicate a tendency for children of similar genders to cluster together. This is in contrast to age (Fig. 2 right), which does not exhibit the same groupings. This could indicate that gendered behavioral differences, characterized by movement and vocal biomarkers, are more distinct than age-specific behavioral differences during the ChAMP mood-induction tasks.

Similarly, the SOM representations of Fig. 3 indicate that individuals with any ADHD, anxiety, or depressive symptoms (regardless of subsequent diagnosis) are nearly segregated completely by U-matrix boundaries from children without reported symptoms. While preliminary, this finding suggests that mood induction task behaviors may indicate the presence of underlying mental health conditions.

**Figure 3.**
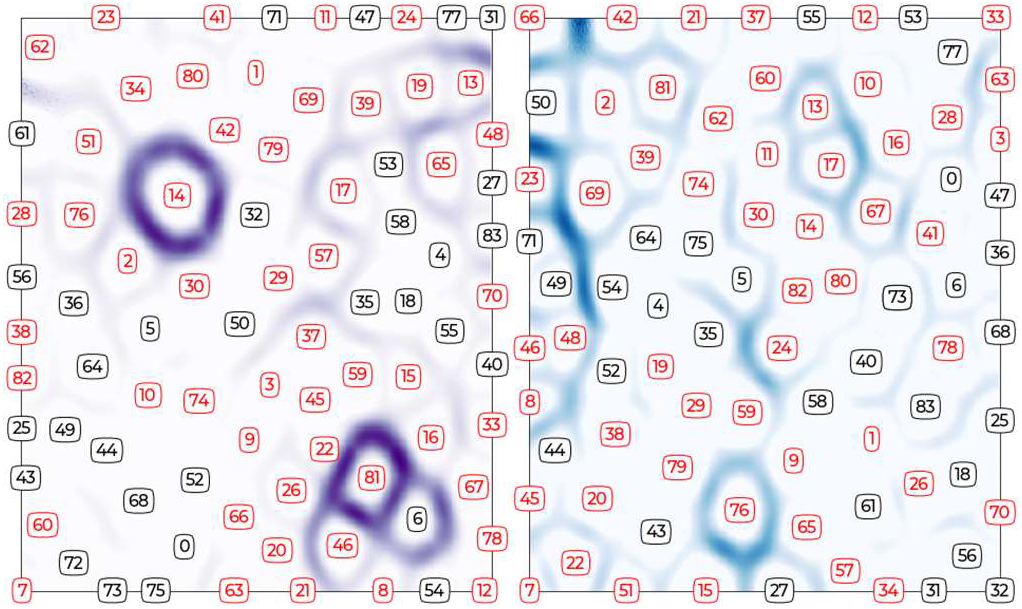
SOMs of the Bubbles (purple, **left**) and Speech (blue, **right**) tasks, encoded by symptom presence (red-at least one symptom of ADHD, anxiety, or depression, black-zero symptoms).

Unlike children with depression (bottom left) or anxiety (top right), the SOM representations of Fig. 4 indicate that those with ADHD (top left) tended to cluster together. Interestingly, these groupings did not persist through ADHD subtypes (e.g., hyperactive, inattentive), which could indicate that the overarching diagnosis may present similarities in behavioral biomarkers, while the specific sub-diagnosis may not be reflected as clearly in these data. However, diagnoses do seem to inhabit relatively unique areas within the SOM such that children with only depression (i.e., no comorbidities) are distant from those with only anxiety and only ADHD (Fig. 4, bottom right). While closer in proximity, there is also very little overlap between those with only anxiety and only ADHD.

**Figure 4.**
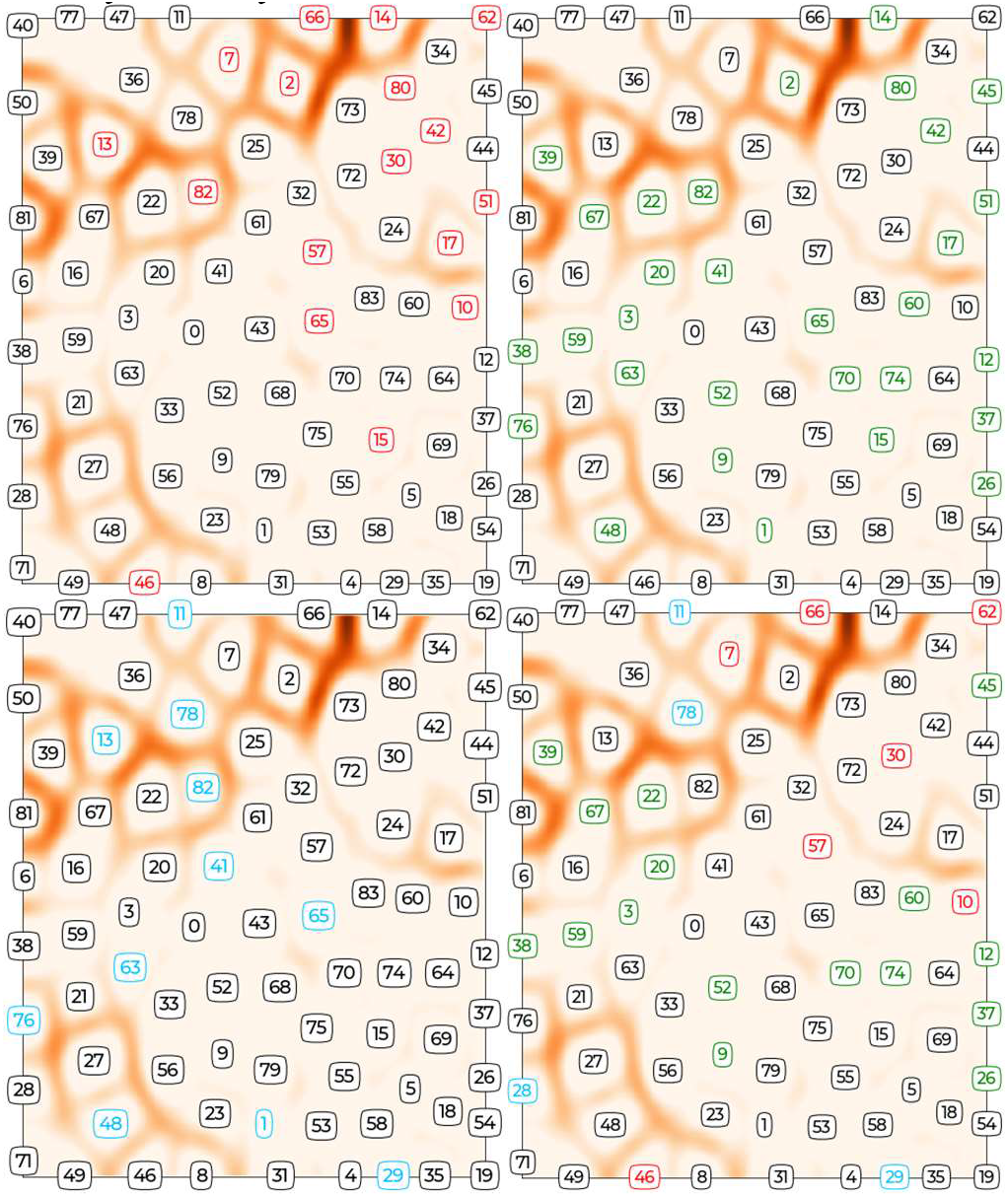
Cross-task SOMs divided and encoded by diagnostic status (top left-ADHD: red, top right-Anxiety: green, bottom left-depression: blue, bottom right-pure anxiety: green, pure depression: blue, and pure ADHD: red). Binary depictions are encoded by status and include comorbid diagnoses (black-no diagnosis, red-diagnosis). Bottom right black-no diagnosis or a comorbid diagnosis.

When examining outliers across task-specific SOMs, we found several interesting features of distinct children (*i*.*e*. children in solo clusters with distinct boundaries separating their behavioral signature from other children). Case studies of these behaviorally distinct children can offer an additional viewpoint as to why specific children may not be segregating as consistently as other children in the study. In the Bubbles task (Fig 3, left) two children were most unique across the trained SOM: child 14 and 81. When assessing factors that may have influenced their behavioral individuality, we found that child 81 was the youngest child (4 years and 1 month) across the study population with a difference of two months to the next oldest child. Child 14, when assessed, was over the 95^th^ percentile for symptom count total, and was diagnosed with two diagnoses. The child was also in the upper percentiles of both of the diagnosis-specific symptom count distributions (98^th^ percentile Anxiety, 95^th^ percentile ADHD). Across both the Approach and Speech Task, child 66 showed unique behavioral signatures in the SOMs. This child had a diagnosis of Combined ADHD and was in the 98^th^ percentile for ADHD symptom count, as well as the 93^rd^ percentile for symptom count overall. These features appeared to be trends for behaviorally distinct children across tasks, as others tended towards higher percentiles for total or diagnostic specific symptom counts (>80^th^ percentile, Approach task: 7, 78, 2, 66; Speech Task: 76, 66; Bubbles task: 14,81) and several others had diagnoses of ADHD-Combined (Approach task: 7, 66, and 2; Bubbles task: 14; Speech task: 66). However, we could not determine patterns for all behaviorally distinct children as others experienced fewer symptoms (<80^th^ percentile, Approach task: 11; Speech task: 23), or no diagnoses (Approach task: 25; Speech task: 23; Bubbles task: 81). Considering factors beyond symptoms and disorders, and their interactions with demographics, is important for future modelling efforts.

### B. Predictive Modeling of Childhood Mental Health Status

Building on these unsupervised clustering results, we now consider results from our supervised learning which aimed to build models for detecting children with Anxiety, Depression, or ADHD. The top performing two models for each diagnosis (Anxiety, Depression, ADHD) are reported in Fig. 5. Models trained to detect ADHD outperformed those for Anxiety and Depression, achieving ROC AUC scores of .65-.72 and correctly identified between 41-71% of children with disorders in the test set. While these results are promising, significant future work is needed to engineer additional theory-driven features, integrate other physiological signals, and continue to explore modeling approaches to optimize performance.

**Figure 5.**
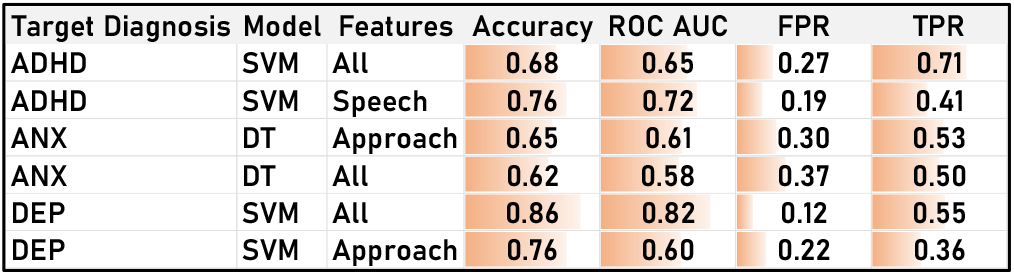
Top 2 highest performing models for each diagnosis (ANX = Anxiety, DEP = Depression, ADHD = ADHD). FPR = false positive rate. TPR = true positive rate.

## IV. Discussion

In this study, we present unsupervised clustering and supervised modeling efforts on a cohort of 84 children, 4-8 years old. We support, via Kohonen SOMs, the need for data collected across diverse populations to fully represent a digital phenotype of childhood mental health status. To the best of our knowledge, prior work has not examined unsupervised clustering of early childhood behavioral task biomarker data. Preliminary findings suggest clustering of ChAMP data may not be best reflected by traditional diagnostic statuses. Instead, impairment, diagnostic subtypes, and perhaps their interaction with demographic factors should also be considered for this data and age range. Examining an unsupervised view of how the feature space is clustering can further inform feature selection and improvements in supervised learning results in future work, especially with the incorporation of features from other, non-smartphone-based wearables. To that end, we have extended our past work by implementing a variety of supervised classification models, identifying consistencies in model performance based on previous work, and specified areas of improvement for future modeling efforts. Specifically, inclusion of additional wearable sensors, subjects, parameter tuning, and modeling approaches (e.g., deep learning).

## V. Conclusion

Our current analyses impact childhood mental health modeling by exploring new methodologies for evaluating biomarker datasets and creating new model benchmarks for this vulnerable population. We leverage past work of biomarker discovery and isolated behavioral task analyses to continue to assess and support our open-access data collection tool, ChAMP.

## Data Availability

A subset of the data produced in this study will be made available at a future date.

## Acknowledgments

This work would not be possible without the wonderful participating families and support of the undergraduate research assistants conducting lab visits.

## Notes

* Research supported by the United States National Science Foundation (NSF Award #2046440) and National Institutes of Health (NIH Award MH123031)

### Competing Interest Statement

The authors have declared no competing interest.

### Funding Statement

Research supported by the United States National Science Foundation (NSF Award #2046440) and National Institutes of Health (NIH Award MH123031)

### Author Declarations

University of Vermont IRB provided ethical approval for this work.

